# Bleeding And Thrombosis In Patients With Out Of Hospital VT/VF Arrest Treated With Extracorporeal Cardiopulmonary Resuscitation

**DOI:** 10.1101/2023.12.07.23299701

**Authors:** Alejandra Gutierrez, Rajat Kalra, Kevin Y. Chang, Marie E. Steiner, Alexandra M. Marquez, Tamas Alexy, Andrea M. Elliott, Meagan Nowariak, Demetris Yannopoulos, Jason A Bartos

**Author notes:** Corresponding author Alejandra Gutierrez MD Cardiovascular Division University of Minnesota 420 Delaware Street SE MMC 508 Minneapolis, MN 55455.

## Abstract

**Background:** Extracorporeal cardiopulmonary resuscitation (eCPR) improves outcomes after out of hospital cardiac arrest (OHCA). However, bleeding and thrombosis are common complications. The post cardiac arrest syndrome including trauma and altered hemostasis creates challenges when balancing the risk of bleeding versus thrombosis. We aimed to describe the incidence and predictors of bleeding and thrombosis and their association with in-hospital mortality.

**Methods:** Consecutive patients presenting with refractory VT/VF OHCA between December 2015 and March 2022 who met criteria for eCPR initiation at our center were included. Patients were anticoagulated with unfractionated heparin while on ECMO. Major bleeding was defined by the extracorporeal life support organization’s (ELSO) criteria as clinically significant bleed associated with transfusion of ≥2 units of red blood cells in 24h or bleeding in a critical area. Thrombosis was defined by clinical signs and symptoms and or imaging evidence of thrombi. Adjusted analyses were done to seek out risk factors for bleeding and thrombosis and evaluate their association with mortality.

**Results:** Major bleeding occurred in 135/200 patients (67.5%) with traumatic bleeding from CPR in 73/200 (36.5%). Baseline demographics and arrest characteristics were similar between groups. In multivariable regression analysis only fibrinogen was independently associated with bleeding (aHR 0.98 per every 10mg/dl rise, 95% CI: 0.96-0.99). Patients who died had a higher rate of bleeds per day (0.21 vs 0.03,p<0.001) though bleeding was not significantly associated with in-hospital death (aHR 0.81, 95% CI: 0.55-1.19). A thrombotic event occurred in 23.5% (47/200). Venous thromboembolism (VTE) occurred in 11% (22/200) and arterial thrombi in 15.5% (31/200). Clinical characteristics were comparable between groups. In adjusted analyses, antithrombin III level on admission (aHR 0.97, 95% CI: 0.94-0.99) and use of dual anti-platelet therapy (DAPT) (aHR 0.31, 95% CI: 0.11-0.86) were associated with the incidence of thrombosis. Thrombosis was not associated with in-hospital death (aHR 0.65, 95% CI: 0.42- 1.03).

**Conclusion:** Bleeding is a frequent complication of eCPR that is associated with decreased fibrinogen levels on admission. Thrombosis is less common occurring in 24% of the cohort. Neither bleeding nor thrombosis was significantly associated with in-hospital mortality.

**Clinical Perspective:** *What is new?:*

- In this large single center study with a protocolized approach to anticoagulation for patients with VT/VF OHCA bleeding as defined by ELSO major bleeding was common occurring in 68% of the cohort while thrombosis was less frequent occurring in 24% with the more than half of the events being arterial thrombi.
- Bleeding events are multifactorial including trauma from prolonged CPR, access site bleeding, and mucosal bleeding.
- Neither bleeding nor thrombosis is associated with overall in-hospital mortality.

*What are the Clinical implications?:*

- The high incidence of bleeding and low incidence of thrombosis with the current approach to anticoagulation which often involves antithrombotic therapy reinforces the need for a better method to risk stratify patients to better tailor anticoagulation strategies.
- The lack of association of bleeding and thrombotic events with mortality support a less aggressive anticoagulation strategy to minimize bleeding.

## Introduction

Extracorporeal cardiopulmonary resuscitation (eCPR) improves survival after out-of-hospital cardiac arrest (OHCA)(1–5). Incidence of EMS-assessed OHCA is estimated to be 350,000 in 2015 per AHA with 7% surviving with a good neurologic outcome(6). Mortality after OHCA has remained poor with further worsening during the COVID 19 pandemic(6). The ARREST trial demonstrated that eCPR, including emergent cannulation with extracorporeal membrane oxygenation (ECMO), improved the absolute survival to hospital discharge by 36% for patients with OHCA due to ventricular tachycardia or ventricular fibrillation (VT/VF) as the presenting rhythm(1). Anticoagulation management of patients presenting with VT/VF OHCA supported by eCPR is challenging with competing thrombogenic insults and high bleeding risk due to trauma from prolonged CPR and coagulopathy related to cardiac arrest and the ECMO circuit. Bleeding is the most common complication of eCPR occurring in 8 to 70% of cases depending on the case series and definition of bleeding used(2, 7, 8) while thrombotic rates are not well documented in the literature.

The bleeding risk associated with eCPR is multifactorial and its quantification is difficult. The contact of blood with artificial surfaces and the sheer stress of the mechanical circuit leads to an acquired coagulopathy by activating the coagulation cascade. Therefore, patients are anticoagulated to prevent thromboembolic events and oxygenator thrombosis(9). eCPR in the setting of severe coronary artery disease presents further challenges as patients often require dual antiplatelet therapy (DAPT) to protect the newly placed coronary stents (8, 10). Moreover, the trauma associated with prolonged CPR and the inflammation and coagulopathy associated with ischemia and reperfusion injury which hallmarks the post cardiac arrest syndrome enhances the already heightened risk of bleeding and thrombosis(11). Data regarding the incidence and predictors of bleeding and thrombosis as well as their impact on outcomes in VT/VF patients treated with eCPR is lacking.

We aimed to describe the incidence, severity, and predictors of bleeding and thrombosis and their association with in-hospital mortality.

## Materials and Methods

### 2.1 Study population

Consecutive patients enrolled in the University of Minnesota (UMN) out-of-hospital refractory VT/VF arrest protocol between December 2015 and March 2022 were eligible for inclusion. Procedures were followed in accordance with the ethical standards of the IRB and in accordance with the Helsinki Declaration of 1975. The study was reviewed and approved by the Institutional Review Board (IRB) at the UMN (IRB #1703M11301). A waiver of informed consent was granted given the research was retrospective involving no more than minimal risk.

The UMN protocol has been previously described (3, 8, 12–16). Briefly, patients were screened by paramedics (EMS) to include adults 18-75 years of age, with a shockable presenting rhythm and no return of spontaneous circulation (ROSC) after 3 shocks or shock-triggered conversion to pulseless electrical activity or asystole, ongoing CPR with a Lund University Cardiac Arrest System (LUCAS^TM^), and an estimated transfer time to the closest cannulation site of less than 30 minutes. At the cannulation site patients were assessed for the presence of the following physiologic criteria: 1) end-tidal CO_2_ >10mmHg; 2) PaO_2_≥50mmHg; 3) lactic acid ≤18mmol/L. Patients meeting the physiologic criteria with ongoing arrest and those who obtained ROSC but were unstable with refractory cardiogenic shock and hypotension despite other interventions such as intra-aortic balloon pump placement (IABP; Maquet Cardiovascular, Wayne, NJ, U.S.A.), vasopressors or inotropes underwent ultrasound-guided and fluoroscopy assisted percutaneous V-A ECMO cannulation. Arterial (15-17Fr) and venous (25Fr) cannulas were typically inserted in the ipsilateral common femoral artery and femoral vein, though contralateral cannulation was used as needed. An antegrade perfusion cannula was inserted into the superficial femoral artery ipsilateral to the arterial ECMO cannula to perfuse the distal lower extremity. Per protocol, a heparin bolus was administered at the time of cannulation(17). All patients underwent coronary angiography with percutaneous coronary intervention (PCI) as indicated following cannulation. An intravascular cooling catheter was placed for targeted temperature management with a goal temperature of 34°C for 24 hours unless uncontrolled bleeding necessitated a higher temperature target. The addition of an IABP was left to the discretion of the interventional cardiologist to maintain coronary perfusion in the setting of severe coronary artery disease or lack of cardiac pulsatility defined as an arterial pulse pressure of less than 10 mmHg after initiation of V-A ECMO and PCI. All patients had non-contrast computerized tomography (CT) of their head, chest, abdomen and pelvis upon admission given the high prevalence of trauma associated with CPR(2, 18).

Patients were admitted to a centralized cardiac intensive care unit (CICU). A continuous unfractionated heparin infusion was administered targeting an ACT goal of 180-200sec.

Anticoagulation targets were adjusted by the treating physician in patients with uncontrolled bleeding. A heparin infusion of 2 U/mL in normal saline was administered at 3mL/hr continuously through the antegrade perfusion cannula to maintain its patency. This infusion continued regardless of the presence of bleeding.

### 2.2 Study definitions and outcome variables

Data was collected by retrospective chart review of medical records. The primary outcome was in-hospital mortality. Secondary outcomes were major bleeding and thrombosis. Major bleeding was defined by the ELSO criteria including clinically overt bleeding associated with a hemoglobin fall of ≥2g/dl or ≥2 units of red blood cells transfused during a 24 hour period(17). This included bleeding in a critical area defined as bleeding occurring in the retroperitoneum, pulmonary, central nervous system, or requiring surgical intervention regardless of the decline of hemoglobin or units of blood transfused. Given initial volume expansion, hemolysis, intra-procedural blood loss, and frequent lab draws, it is common for patients to drop their hemoglobin values significantly with successful V-A ECMO resuscitation after cardiac arrest. Therefore, we scrutinized the medical records to identify clinical signs of bleeding noted by the treatment team or present on imaging associated with transfusions or hemoglobin drop to determine major bleeding events. A declining hemoglobin level without clinical bleeding was not considered a bleeding event.

The presence of thrombosis was determined by imaging studies with corresponding clinical correlates of venous or arterial thrombi. This was further classified into: 1 arterial thrombi including stroke as evidenced by imaging, thrombi in the left atrium, left ventricle, aortic valve or other arterial bed such as the superior mesenteric artery and oxygenator thrombosis requiring exchange of the oxygenator and 2. Venous thrombi including DVT and IVC thrombosis, right atrial thrombi or pulmonary embolism.

The first arterial blood gas collected just prior to ECMO cannulation was used to establish initial hemoglobin level. Other laboratory parameters were obtained from the first complete set of blood work performed on arrival to the CICU.

### 2.3 Statistical Analysis

Continuous variables were represented as means with standard deviations and compared using the t-test or analysis of variance (ANOVA) tests. Non-normal continuous variables were represented as medians with interquartile ranges and compared with the Wilcox Rank-Sum test. Categorical variables were represented as counts with proportions and compared with the chi-squared test.

To identify predictors of major bleeding, the association of age, CPR time, baseline use of anticoagulation, use of DAPT during the admission, INR, fibrinogen, antithrombin III and platelet count were first evaluated in unadjusted analyses. These factors were selected due to the established data suggesting their association with bleeding and thrombosis. Predictors that were significant at p <0.20 in unadjusted analyses were included via stepwise selection in an adjusted (multivariable) Cox proportional hazards model to determine which factors remained significant predictors of major bleeding events. The same method was used to determine which clinical factors were significant predictors of thrombotic events.

Cox multivariable regression analyses were done to account for the effect of clinically important covariates in the association between major bleeding and thrombosis with in-hospital death. Age, sex, CPR time, use of DAPT were chosen *a priori* as covariates in Cox multivariable regression analyses. These variables were chosen due to their acknowledged associations with bleeding and thrombotic events. The proportional hazard assumption was checked using Schoenfeld’s residuals. To overcome immortal time bias for VTE, landmark analysis was performed using 5 days as the landmark time for this survival analysis(19). For the survival analyses that assessed the association of major bleeding events with in-hospital death, major thrombosis was also included as a covariate in the multivariable analyses, due to the tendency of thrombosis to affect clinical anticoagulation management. Similarly, major bleeding events were included as a covariate in the survival analyses that assessed the association of thrombosis with in-hospital death.

In-hospital death may be a competing event for the occurrence of major bleeding events and/or thrombosis during treatment with VA-ECMO. Thus, competing risk analyses were performed to evaluate factors that may be associated with major bleeding events and thrombosis, respectively. The cumulative incidence of major bleeding events and thrombosis was estimated using Gray’s test. In-hospital death due to causes other than bleeding events or thrombosis were the competing events. The results were represented as sub-hazard ratios (sHRs) with 95% confidence intervals. The Fine-Gray model was used to perform multivariable regression. The level of statistical significance was set at <0.05 for all analyses. The analyses were done with IBM SPSS v28 (Chicago, IL, U.S.A.) and StataMP 17.0 (College Station, TX, U.S.A.).

## Results

### 3.1 Study population

A total of 260 patients presented within criteria with a VT/VF cardiac arrest and achieved an organized rhythm at the end of the cardiac catheterization case. 23% of patients achieved stable ROSC and did not require V-A ECMO to stabilize their cardiogenic shock and were therefore excluded from this analysis. The final study population consisted of 200 patients.

Baseline characteristics are shown in **Table 1**. The majority of the cohort was male (81%) with a mean age of 57 (12.2) years. Comorbid conditions included coronary artery disease in 26% (52/200), congestive heart failure (CHF) in 12.5% (25/200), diabetes in 24.5% (49/200) and hypertension in 35.5% (71/200). Prior to admission medications included: a direct oral anticoagulant in 6/200 (3%), a vitamin K antagonist in 4/200 (2%), aspirin in 29/200(14.5%) and a P2Y12 inhibitor in 3/200 (1.5%). The average CPR time from 911 call or identified arrest by EMS to V-A ECMO initiation was 62± 19 minutes. An IABP was placed in 71% (142/200) of the cohort. Revascularization was performed in 121/200 (60.5%) of patients and DAPT was given to 62.0% (124/200) of the cohort.

**Table 1.** Demographics, baseline and arrest characteristics of patients with and without major bleeding and with and without a thrombotic event.

|  | Overall Cohort (N=200) | No major bleeding (N=65) | Major bleeding (N=135) | P value | No thrombotic events (N=153) | Thrombotic events (N=47) | P value |
| --- | --- | --- | --- | --- | --- | --- | --- |
| *Baseline Characteristics* |  |  |  |  |  |  |  |
| Gender, Male | 162(81.0) | 54(83.1) | 108(80.0) | 0.74 | 122(79.7) | 40(85.1) | 0.54 |
| Age, years | 57.0(12.2) | 55.5(12.6) | 57.7(11.9) | 0.24 | 56.8(12.1) | 57.8(12.5) | 0.61 |
| BMI, Kg/m<sup>2</sup> | 30.7(6.2) | 30.2(25.9,35.4) | 29.7(26.8,33.8) | 0.73 | 29.7(26.8,33.7) | 30.6(25.9,33.9) | 0.99 |
| *Comorbidities* |  |  |  |  |  |  |  |
| Coronary artery disease | 52(26.0) | 14(21.5) | 38(28.1) | 0.41 | 38(24.8) | 14(29.8) | 0.62 |
| Congestive Heart Failure | 25(12.5) | 6(9.2) | 19(14.1) | 0.48 | 18(11.8) | 7(14.9) | 0.77 |
| Diabetes mellitus | 49(24.5) | 14(21.5) | 35(25.9) | 0.62 | 35(22.9) | 14(29.8) | 0.44 |
| End stage renal disease | 4(2.0) | 1(1.5) | 3(2.2) | 1.00 | 3(2.0) | 1(2.1) | 1.00 |
| Hyperlipidemia | 49(24.5) | 13(20.0) | 36(26.7) | 0.39 | 35(22.9) | 14(29.8) | 0.44 |
| Hypertension | 71(35.5) | 17(26.2) | 54(40.0) | 0.08 | 49(32.0) | 22(46.8) | 0.09 |
| *Arrest characteristics and treatments prior to ICU admission* |  |  |  |  |  |  |  |
| CPR time, min | 62(19) | 61(19) | 63(20) | 0.84 | 62(20) | 61(17) | 0.69 |
| IABP | 142(71.0) | 42(64.4) | 100(74.1) | 0.23 | 105(68.6) | 37(78.7) | 0.25 |
| Heparin bolus during cannulation and PCI, Units | 12578(4353) | 12585(5202) | 12575(3901) | 0.99 | 12544(4588) | 12682(3588) | 0.85 |
| Cangrelor | 16(8.0) | 3(4.6) | 13(9.6) | 0.34 | 11(7.2) | 5(10.6) | 0.65 |
| GP iib/iiia inhibitor | 6(3.0) | 2(3.1) | 4(3.0) | 1.000 | 3(2.0) | 3(6.4) | 0.29 |
| DAPT | 124(62.0) | 33(50.8) | 91(67.4) | 0.03 | 90(58.8) | 34(72.3) | 0.13 |
| Ticagrelor | 121(60.5) | 33(50.8) | 88(65.2) | 0.11 | 87(56.9) | 34(72.3) | 0.08 |
| Clopidogrel | 3(1.5) | 0(0.0) | 3(2.2) | 0.55 | 3(2.0) | 0(0.0) | 0.81 |

BMI, body mass index; CPR, cardiopulmonary resuscitation; DAPT, dual antiplatelet therapy; IABP, intra-aortic balloon pump;

The hemoglobin on initial presentation to the emergency department or catheterization lab was 12.6±2.6 g/dl. Other lab parameters were obtained at the time of arrival to the CICU. Most blood tests were above normal limits (**Figure 1**) including: ALT (187±199 U/L), AST (372±410 U/L), INR(2.0±0.9), ACT (224±63), PTT (195±72sec) and D-dimer(16±7Ug/mL).

**Figure 1.**
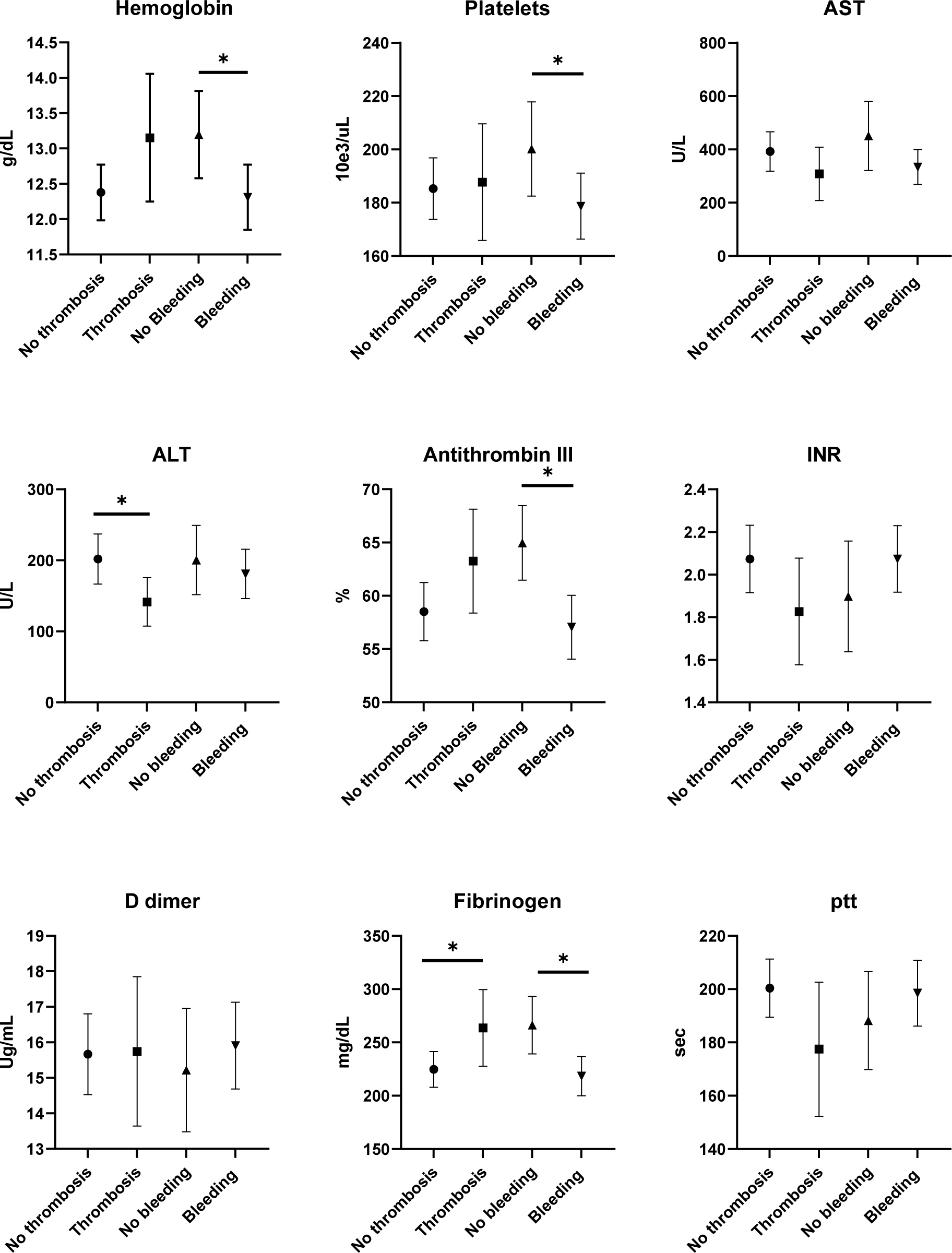
Hematologic markers on arrival to the intensive care unit stratified by patients with and without thrombosis and those with and without major bleeding.

Antithrombin III levels were often below the normal range of our laboratory (>85%) with an average of 59.6% ± 16.9 while the average fibrinogen of our cohort was within normal limits at 234±109 mg/dL.

### 3.2 Predictors of Bleeding

Major bleeding was common occurring in 135/200 patients (67.5%). There was more than one site of bleeding in 91/200 (45.5%) patients. Minor bleeding which did not meet criteria for ELSO major bleeding occurred in 21/200(10.5%) of patients and there was no clinical evidence of bleeding in 44/200 (22%) patients. Details regarding major bleeding site are shown in **Figure2A**. The most common site of major bleeding was access site bleeding. This was present in 106/200 (53.0%) with 6 patients requiring a surgical intervention to repair this.

Intracranial bleeding (ICH) occurred in 10/200 (5.0%) including 3 cases of subarachnoid hemorrhage, 2 intraventricular hemorrhages, and 5 intraparenchymal hemorrhages. The median day of diagnosis of ICH was 3 (1,7) days. CPR-related trauma was frequent, occurring in 73/200 (36.5%) of the cohort including mediastinal hematoma in 51/200(26%), 14/200 (7%) with pericardial bleeding, 11% (22/200) with a hemothorax and 7% (14/200) intraperitoneal bleeding. Gastrointestinal bleeding was clinically diagnosed in 33/200 (16.5%) patients. Most cases were managed medically with only 6/200 cases (4.7%) undergoing upper endoscopy. The majority of bleeding events (92%) happened while the patients were on V-A ECMO. The first major bleeding event was frequently within the first days of hospitalization, with a median time from admission to bleeding of 1 (0,2) day with 50% of patients bleeding on day 0-1 of admission and 74% of all bleeding events occurring before the third day of the hospitalization (**Figure 2B**) .

**Figure 2.**
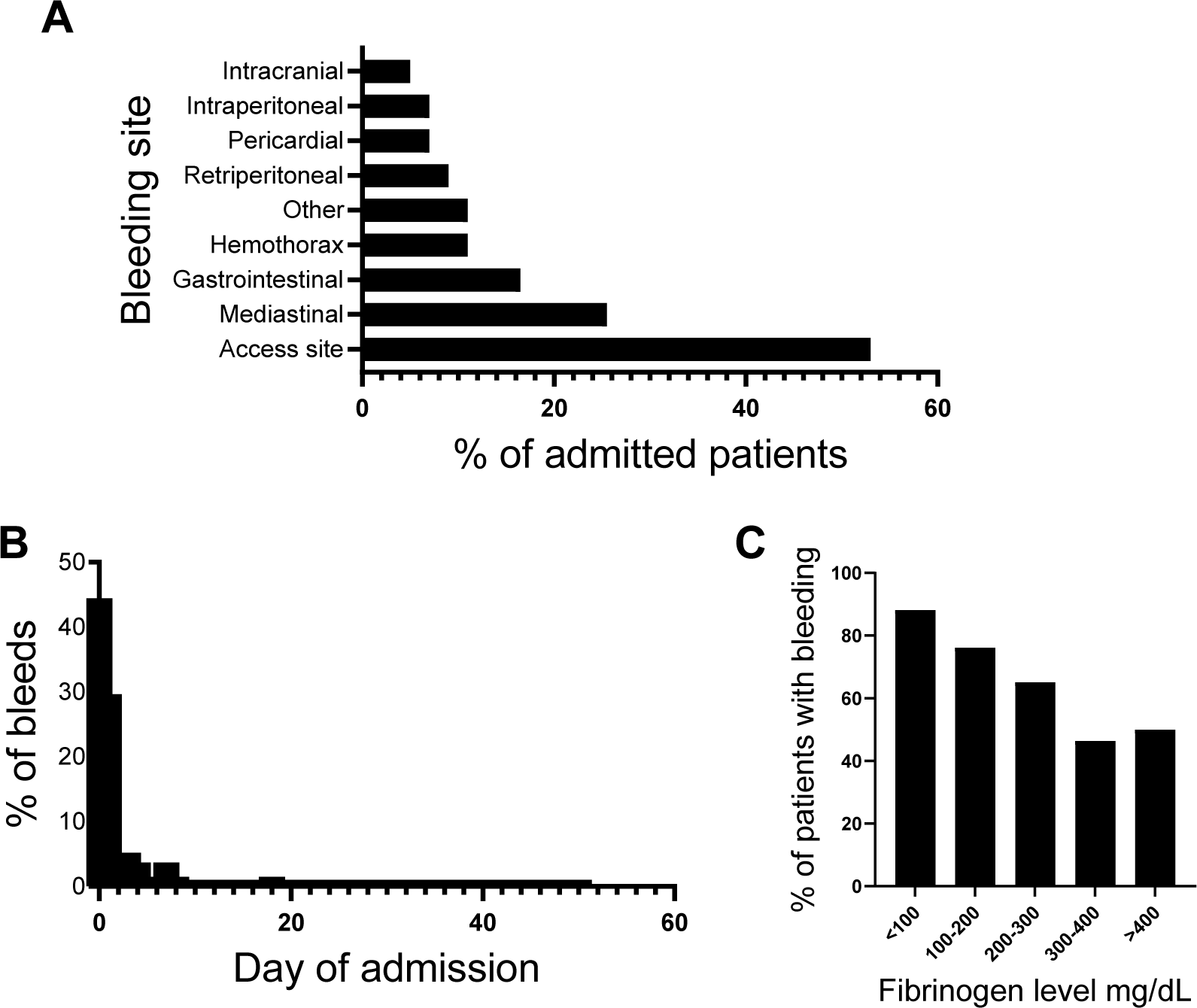
Characteristics of major bleeding events. A. Sites of major bleeding. B. Percentage of admitted patients who had a major bleeding event. C. Percentage of patients who had a major bleeding event depending on admission fibrinogen level.

Baseline comorbidities were comparable between those who had major bleeding and those without major bleeding (**Table 1**). In unadjusted analysis, those with a bleeding event had a lower hemoglobin (12.3 vs 13.1g/dL; p=0.05), platelets (179 vs 200 x 10e3/uL, p=0.04), fibrinogen (218 vs 266 mg/dL, p=0.004), and antithrombin III levels (57 vs 65%, p<0.001) on admission (**Figure 1**). Our patients had an average D-dimer of 15.9(7.2) vs 15.2(7.0) ug/mL for bleeding and non-bleeding patients respectively with a normal for our laboratory being 0.0- 0.5ug/mL and no significant difference among groups (p=0.52).

Use of procedural anticoagulation and antiplatelet medications were not associated with bleeds. The use of iib/iiia inhibitors and cangrelor was uncommon (6/200(3.0%) and 16/200(8.0%), respectively). The average dose of heparin given for ECMO cannulation and rate of PCI was not significantly different between groups.

In unadjusted analyses age, antithrombin III, and fibrinogen met the prespecified p <0.20 threshold for inclusion in the adjusted model. In adjusted analyses, only fibrinogen was associated with incident major bleeding events with every 10 unit rise in fibrinogen associated with 0.98 times the hazard of major bleeding (HR 0.98, 95% CI: 0.95-0.99) (**Central Figure)**.

The risk of bleeding seemed to heighten with fibrinogen levels below 300mg/dL (with 47.7% of patients having a major bleed in those with a fibrinogen > 300mg/dL vs 72.8% in those with a fibrinogen <300mg/dL, p=0.003) **(Figure 2C).**

### 3.3 Outcomes associated with bleeding

In unadjusted analyses, the occurrence of a major bleeding event was not significantly associated with in-hospital death. Similarly, after adjustment for clinically relevant factors, having an ELSO major bleeding event was not associated with in-hospital death (aHR 0.81, 95% CI: 0.55-1.19).

Hospital course and outcomes are shown in **Table 2**. In unadjusted analysis, major bleeding was associated with higher transfusion needs with patients who had a bleeding event receiving a median of 7(4,12) units of red blood cells, 2(0,5) units of platelets and 1(0,3) unit of fresh frozen plasma. Patients who had a bleeding event had similar rates of renal replacement therapy(30.4 vs 21.5%; p=0.25), extubation (29.6 v 20.0%; p=0.20) and successful V-A ECMO decannulation (48.1 vs 32.3%;p=0.19). For those patients who survived to extubation, the length of mechanical ventilation was longer in patients who had a major bleeding event (9 vs 6 days, p=0.05). The median length of hospital stay was 8 (2,21) days. Having a bleeding event was associated with a longer length of hospitalization with a median of 10(3,22) compared to 4(2,14) (p=0.01)days of admission.

**Table 2.** Treatment characteristics and hospital course of patients who had a bleeding event and those without bleeding events.

|  | Overall cohort<br>(N=200) | No bleeding<br>(N=65) | Major bleeding<br>(N=135) | P value |
| --- | --- | --- | --- | --- |
| *Transfusions during the hospitalization* |  |  |  |  |
| Red blood cells, units | 4.5(1.0,10.0) | 1.0(0.0,2.0) | 7.0(4.0,12.0) | **<0.001** |
| Platelets, units | 1(0.0,4.0) | 0.0(0.0,1.0) | 2.0(0.0,5.0) | **<0.001** |
| Fresh Frozen Plasma, units | 1.0(0.0,3.0) | 0.0(0.0,0.0) | 1.0(0.0,3.0) | **<0.001** |
| *Hospital Course* |  |  |  |  |
| Renal Replacement Therapy, N(%) | 55(27.5) | 14(21.5) | 41(30.4) | 0.25 |
| Days on continuous dialysis*(N=14) | 6.0(4.0,18.0) | 3.5(3.0,4.0) | 8.0(4.0,20.0) | 0.20 |
| Thrombosis, N(%) | 47(23.5) | 11(16.9) | 36(26.7) | 0.16 |
| Extubation, N(%) | 53(26.5) | 13(20.0) | 40(29.6) | 0.20 |
| Days on mechanical ventilation**<br>(N=53), | 8.0(6.0,13.0) | 6.0(5.0,8.0) | 9.0(6.0,14.0) | **0.05** |
| Days on VA ECMO | 4.0(3.0,5.0) | 3.0(2.0,4.0) | 4.0(2.0,5.0) | 0.05 |
| VA ECMO decannulation, N(%) | 95(47.5) | 21(32.3) | 65(48.1) | 0.19 |
| Length of hospital stay, days | 8.0(2.0,20.5) | 4.0(2.0,14.0) | 10.0(3.0,22.0) | **0.01** |
| Survival to hospital discharge, N(%) | 64(32.0) | 17(26.2) | 47(34.8) | 0.29 |

\*calculated for patients who survived to discontinuation of continuous dialysis
\*\*Including patients who survived to liberation from mechanical ventilation

In the adjusted competing risks analyses, age (sHR 1.01, 95% CI: 0.99-1.03), sex (sHR 0.75, 95% CI:0.44-1.27), CPR time (sHR 1.00, 95% CI: 0.99-1.01), use of DAPT during the hospitalization (sHR: 0.96, 95% CI: 0.62-1.49), were not associated with major bleeding or all-cause in-hospital mortality.

### 3.5 Thrombotic events

Arterial or venous thrombotic events were identified in 23.5% (47/200) patients. The sites of thrombi are shown in **Figure 3A**. Venous thromboembolism including deep vein thrombosis (DVT), right atrial thrombi and pulmonary embolism (PE) occurred in 22/200 (11.0%). Arterial thromboembolism was seen in 31/200 (15.5%) with 6 patients experiencing both arterial and venous thrombi during their hospitalization. Oxygenator thrombosis requiring exchange of the membrane was uncommon, occurring in 1.5% of the cohort(3/200). Two of these occurred the day of admission soon after cannulation and the other one occurred 12 days after cannulation. Despite continuous anticoagulation infusion through the reperfusion cannula, problems with thrombus formation in the tubing or catheter occurred in 5 patients (2.5%). Two other patients (1%) had lower limb ischemia requiring thrombectomy to salvage the limb. Thrombosis occurred at a median of 5(1,12) days from the day of admission. Arterial thrombi presented earlier (2 [1,6] days) compared to VTE events (12 days [9,16]) **Figure 3B**.

**Figure 3.**
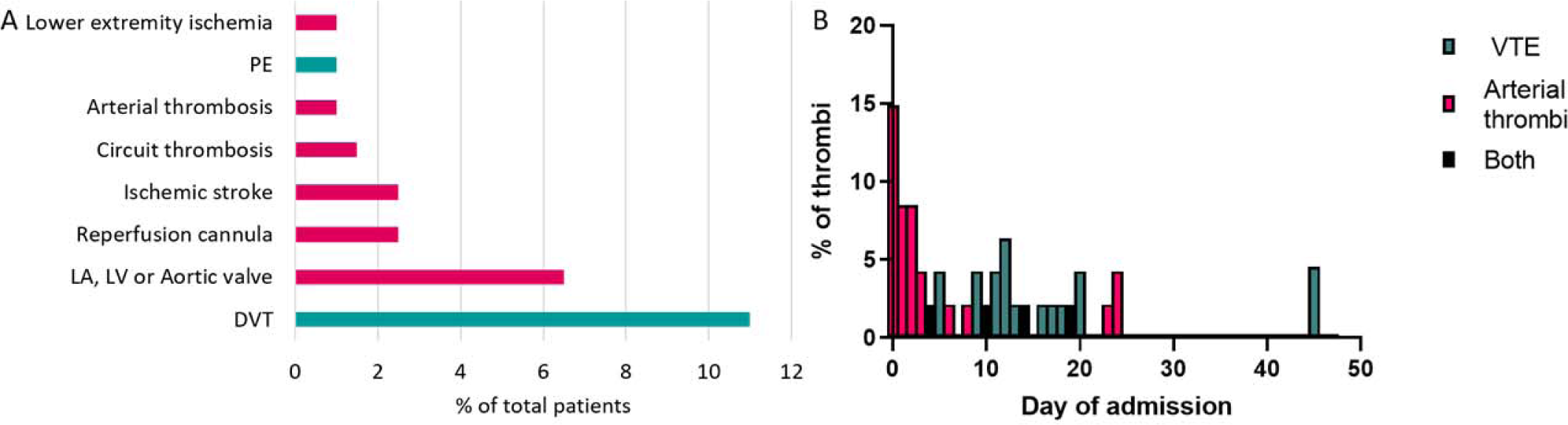
Characteristics of thrombotic events. A. Percentage of patients with different sites of thrombosis with VTE(green) and arterial thrombosis (pink). B. Percentage of admitted patients who present thrombosis per day of admission. * arterial thrombosis refers to aorta and/or SMA thrombosis.

**Figure 4.**
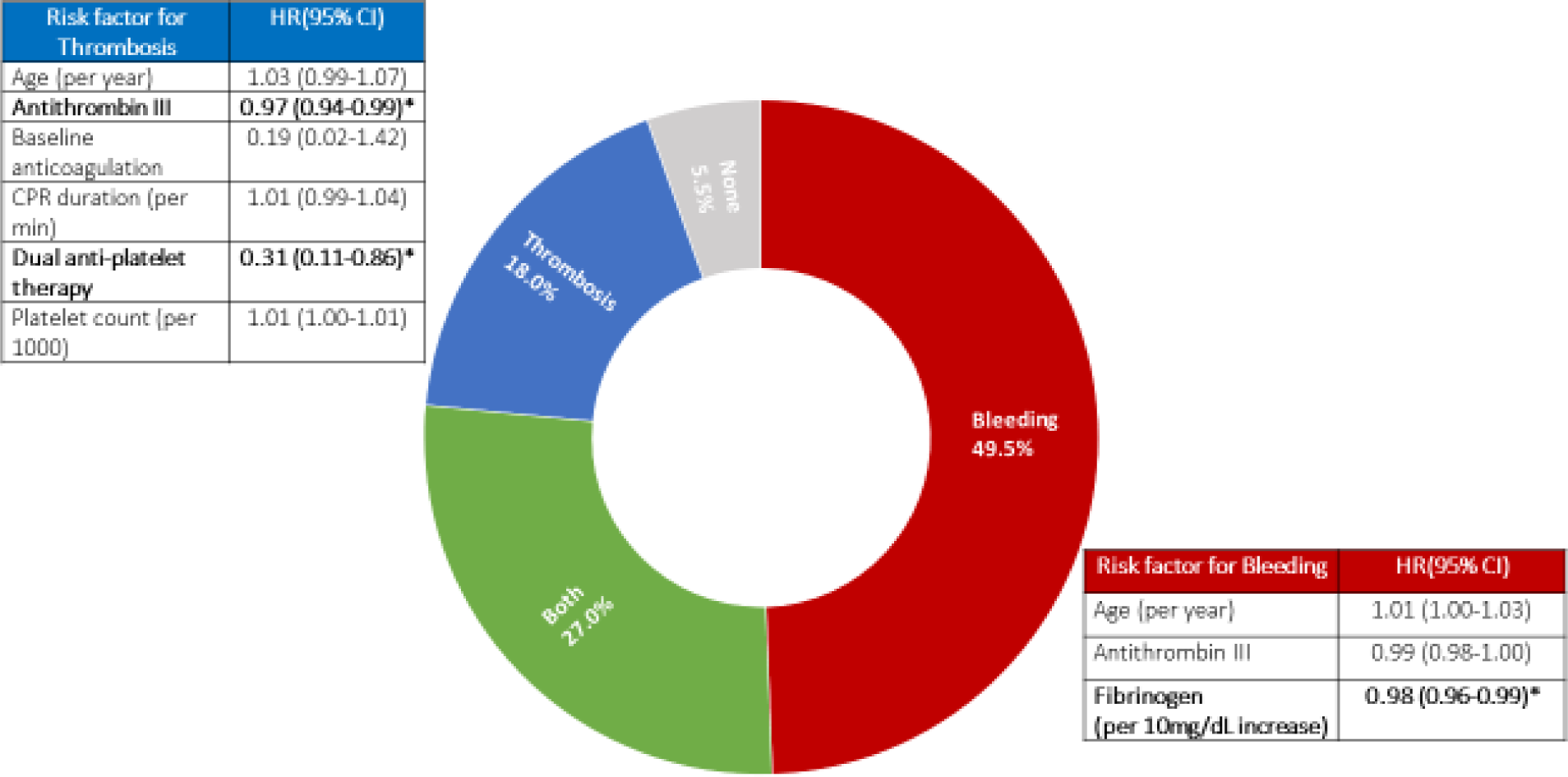
Central figure. Rates of bleeding and thrombosis. Multivariable model of risks of bleeding and thrombosis. * denotes statistical significance.

There were no differences in baseline characteristics, comorbidities or arrest characteristics in the groups with and without thrombosis (**Table 1).** Those who had a thrombotic event had a higher fibrinogen level (264 vs 225, p=0.03) and a lower ALT (141 vs 202, p=U/L) compared to those who did not have a thrombotic event (**Figure 1**). Treatment characteristics of patients with and without thrombosis are shown in **Table 3**. The use of an IABP was not associated with increased thrombotic events. A thrombus in the inferior vena cava was detected in 6/300 (3.0%) accounting for 6/22 (27%) of all the deep venous thromboembolisms. Length of V-A ECMO cannulation was associated with a longer time on V-A ECMO predominantly due to longer runs in patients with arterial thrombi but was not associated with the rate of VTE **(Table 3).** In the univariate analyses, age, CPR time, baseline use of anticoagulation, DAPT administration, antithrombin III, and platelet count met the pre specified p <0.20 threshold for inclusion in the adjusted model. In the adjusted analyses, a higher level of antithrombin III (aHR 0.97, 95% CI: 0.94-0.99) and use of DAPT (aHR 0.31, 95% CI: 0.11-0.86) were associated with a reduced incidence of thrombosis **(Central Figure).**

**Table 3.** Treatment characteristics and hospital course of patients who had a thrombotic event including arterial thrombosis and VTE and those without thrombosis.

|  | No thrombosis<br>(N=153) | Any Thrombosis<br>(N=47) | P value | No arterial thrombosis<br>(N=169) | Arterial thrombosis<br>(N=31) | P value | No VTE<br>(N=178) | VTE<br>(N=22) | P value |
| --- | --- | --- | --- | --- | --- | --- | --- | --- | --- |
| *Transfusions during the hospitalization* |  |  |  |  |  |  |  |  |  |
| Red blood cells, units | **4(1,9)** | **7(2,12)** | **0.01** | **4.0(1.0,9.0)** | **7.0(2.0,12.0)** | **0.05** | 4.0(1.0,9.3) | 7.0(2.8,12.0) | 0.07 |
| Platelets, units | **1.0(0.0,3.0)** | **2.0(0.0,5.0)** | **0.005** | 1.0(0.0,3.0) | 2.0(0.0,6.0) | 0.06 | **1.0(0.0,4.0)** | **2.0(1.0,5.5)** | **0.02** |
| Fresh frozen | 1(0,2) | 0(0,2) | 0.50 | 0(0,2) | 1(1.3) | 0.39 | 1(0,2) | 0(0,1) | 0.09 |
| plasma,<br>units |  |  |  |  |  |  |  |  |  |
| *Hospital course* |  |  |  |  |  |  |  |  |  |
| Renal<br>Replaceme<br>nt Therapy | 37(24.2) | 18(38.3) | 0.09 | 40(23.7) | 15(48.4) | 0.00<br>9 | 49(27.5) | 6(27.3) | 1.00 |
| Days on<br>continuous<br>dialysis*<br>(N=14) | 4.5(2.8,11<br>.3) | 17.0(6.0,22<br>.0) | 0.12 | 5.0(3.0,18<br>.0) | 12.0(4.0,12<br>.0) | 0.39 | 5(3,12) | 22(4,) | 0.18 |
| Major<br>bleeding<br>event | 99(64.7) | 36(76.6) | 0.18 | 111(65.7) | 24(77.4) | 0.28 | 118(66.3) | 17(77.3) | 0.42 |
| Extubation,<br>N(%) | 37(24.2) | 16(34.0) | 0.25 | 48(28.4) | 5(16.1) | 0.23 | **39(21.9)** | **14(63.6)** | **<0.00**<br>**1** |
| Days on<br>mechanical<br>ventilation*<br>* (N=53),<br>N(%) | 7.0(5.5,11<br>.0) | 11.5(6.5,18<br>.5) | 0.07 | 7.5(6.0,13<br>.0) | 13.0(8.0,17<br>.0) | 0.17 | 7.0(6.0,11<br>.0) | 11.5(7.5,15.<br>5) | 0.09 |
| VA ECMO<br>decannulati<br>on, N(%) | 60(39.2) | 26(55.3) | 0.07 | 73(43.1) | 13(41.9) | 1.00 | **68(38.2)** | **18(81.8)** | **<0.00**<br>**1** |
| Length of<br>VA ECMO<br>run days | **3.5(2.4)** | **4.8(3.1)** | **0.008** | **3.6(2.5)** | **4.9(3.1)** | **0.04** | 3.7(2.6) | 4.8(2.9) | 0.12 |
| Length of<br>hospital<br>stay, days | **6(2,18)** | **15(5,29)** | **<0.00**<br>**1** | 8(2,21) | 10(3,22) | 0.37 | **6.0(2.0,17**<br>**.3)** | **28.0(15.0,3**<br>**7.8)** | **<0.00**<br>**1** |
| Survival to<br>hospital<br>discharge,<br>N(%) | 46(30.1) | 18(38.3) | 0.38 | 58(34.3) | 6(19.4) | 0.15 | **49(27.5)** | **15(68.2)** | **<0.00**<br>**1** |

### 3.6 Outcomes associated with thrombosis

Patients who had a thrombotic event had a higher number of transfusions of red blood cells and platelets (7 versus 4 units; p=0.01 and 2 versus 1 units; p=0.005 respectively) (**Table 3**). There was no significant difference in terms of renal replacement therapy or rates of extubation. However, the length of stay was significantly longer for those who had a thrombotic event.

In the unadjusted analyses, the occurrence of a thrombotic event was not associated with in hospital death (**Table 3**). After adjustment for age, sex, CPR time, use of DAPT and the occurrence of major bleeding, the occurrence of thrombosis was not associated with in-hospital death (aHR 0.65, 95% CI: 0.42-1.03). Adjustment for competing risk of death also negated the difference observed in the unadjusted analysis. When stratified by type of thrombosis, VTE was associated with in hospital mortality in unadjusted (**Table 3**) and adjusted analysis (aHR 0.22, 95% CI 0.09-0.60). The rate of VTE was associated with the length of stay with a rate of 0.01+/- 0.023 per day for those who survived compared to 0.003 +/-0.015 for those who died (p=0.008). Each additional day of hospitalization was associated with an OR of 1.05 of VTE (95% CI: 1.0- 1.1, p<0.001) making it very sensitive to survival bias. In a landmark analysis, excluding patients who died before the first 5 days of admission (none of whom had a VTE), VTE was no longer associated with in hospital mortality (HR 0.44, 95% CI: 0.17-1.12).

Arterial site thrombosis was not associated with in hospital death in unadjusted (**Table 3**) or adjusted analysis (aHR1.2, 95% CI 0.73-1.94). In the adjusted competing risks analyses, only age was associated with increased arterial site thrombosis (sHR 1.04, 95% CI 1.00-1.09).

## Discussion

In this large observational single center study, we report the incidence and risk factors associated with bleeding and thrombosis and their association with in-hospital mortality. Major bleeding as defined by ELSO is a common complication of eCPR, affecting 67.5% of our cohort. In multivariable analysis, the only risk factor associated with bleeding was decreasing levels of fibrinogen. Thrombosis was less common than bleeding, occurring in 23.5% of patients with 11.0% of patients having a VTE and 15.5% an arterial thrombi. A higher level of antithrombin III on admission and the administration of DAPT was associated with decreased thrombotic risk. Neither bleeding nor thrombosis was associated with in-hospital mortality when results were adjusted for clinically significant factors.

Reported rates of bleeding in the eCPR population vary widely(5, 20–25), but are higher than those of other types of ECMO including V-A ECMO for other cardiac indications (21, 25) or V-V ECMO (21). The heightened risk of bleeding is thought to be related to trauma from prolonged CPR, hypothermia, and coagulopathy associated with the post cardiac arrest syndrome(5, 11, 22). In our cohort, we found an incidence of intracranial hemorrhage of 5% which is similar to that reported in other eCPR cohorts (4.3-5.6%)(21, 23), while 67.5% of patients had major bleeding events. Although elevated, this is concordant with other eCPR cohorts, including the study by Otani et al published in 2018 which reported a 70% (71/233) bleeding rate(22). In their study, major bleeding was assessed by BARC (Bleeding Academic Research Consortium) criteria defined as Type 3 or higher correlating to overt bleeding plus a hemoglobin drop of ≥3g/dL within 24 h of hospital admission(22). Similarly, the CHEER study had a bleeding rate of 70% (18/26) though they do not specify how they defined this outcome(5). However other studies of eCPR patients quote lower bleeding rates including: 38% as defined by the BARC type 3 or higher (20), 39% assessed by BARC type 3b or higher(24), 26% per ELSO criteria in the ELSO registry(25), and 20-22% without a clear definition of major bleeding(23, 26). The discrepancy is likely due to the various definitions of bleeding, with ELSO-defined major bleeding being a more sensitive criteria including a drop of ≥2g/dl/24h(26).

In unadjusted analysis, decreased levels of fibrinogen, antithrombin III and platelet count were associated with increased risk of major bleeding. Prior studies on eCPR patients suggest that hyperfibrinolysis may increase bleeding risk. In a study including 133 patients, higher D-dimer level on admission (18.8ug/mL vs 6.7,ug/mL) was associated with major bleeding(22)..

We did not find significant associations between initial D-dimer and bleeding, but the D-dimer levels of our cohort were very elevated (15.7 ±0.5ug/mL (normal for our laboratory: 0.0- 0.5ug/mL) and frequently reported as >20 ug/mL. This may have led to an underestimation of the actual D-dimer levels in bleeding patients and may explain the disparate findings. However, markers of coagulation factor consumption including low fibrinogen and low antithrombin III were associated with bleeding. In adjusted analysis, decreased level of fibrinogen was the only variable associated with increased bleeding risk. Another study including all V-A ECMO patients identified fibrinogen among other factors to be predictive of bleeding (25). Low platelet count, elevated D-dimer, and low fibrinogen has been observed in other eCPR populations with an elevated disseminated coagulation score (DIC) score being associated with mortality(24, 27).

These studies excluded patients on DAPT whereas we had a significant proportion of patients on DAPT and found a persistent association with markers of consumption and increased bleeding risk.

Major bleeding increased in-hospital morbidity but was not associated with increased mortality. The lack of association with in-hospital mortality diverges from some published ECMO studies where bleeding has been associated with increased mortality(9, 21, 28). In contrast to our data these studies included veno-venous and veno-arterial ECMO support(9, 21, 28) and pediatric and adult patients(9, 21). Further, it is possible that bleeding as defined by the criteria for ELSO major bleeding is an insensitive marker of major events. However, other studies have reported a lack of association between bleeding and in hospital death. In an unadjusted analysis utilizing BARC type 3 or higher by Otani et al, the rates of bleeding were similar in survivors and non-survivors treated with eCPR (46v33%, p=0.44). Similarly in a study by Sahli et al including all-comers requiring ECMO support, development of bleeding complications during ECMO was not an independent predictor of mortality though they did not provide their definition of bleeding(23). Major bleeding was associated with increased transfusion needs in this study in agreement with prior reports in the literature (9, 21, 23, 25) and with longer time on mechanical ventilation.

Thrombotic complications are sparsely reported in the eCPR literature. We found a rate of 22.5% which is similar to that published in another study including eCPR patients with a rate of thrombosis of 21%(20), but higher than the 7% reported in a recent meta-analysis(26). The risk of limb ischemia in the literature is around 16-24%(20, 26) whereas we had a much lower rate of 1%. We routinely place antegrade perfusion catheters at the time of cannulation and infuse heparin through that catheter. Further, our patients were uniformly eCPR patients with an anticoagulation strategy that utilized an unfractionated heparin infusion targeting an ACT of 180- 200 sec. This is not a universal practice and may account for these differences(26). In our study, predictors of thrombosis included lower levels of antithrombin III and patients without DAPT administration. Data regarding risk factors for thrombosis is lacking. In a meta-analysis including all types of ECMO, the anticoagulation strategy was not significantly associated with thrombotic risk though the use of antiplatelet therapy was not reported(26).

In this cohort, thrombosis was not associated with increased mortality. Consistent with other eCPR(20) and general ECMO (26) literature, we did not find any association between survival and thrombosis(20). Thrombotic complications of eCPR include arterial and venous thrombi and their pathophysiology differs significantly. Arterial thrombi typically present in the first few days of admission while patients are highly reliant on the ECMO circuit and the ejection fraction is frequently limited (16). The association of VTE with in-hospital mortality is likely due to immortal time bias, with a rate of thrombosis per day of 0.01 +/- 0.02 for those who survived vs 0.003+/- 0.02 for those who died. VTE is highly dependent on length of hospital stay as has been shown in numerous cohorts including our data where the risk of VTE increased by 5% per each additional day of hospitalization(29, 30). A landmark analysis was performed to overcome this important survival bias. In a study including routine ultrasound evaluation for VTE post decannulation, DVT was found in 15/30 (50%) of those who survived to decannulation where V-V ECLS and length of cannulation were the only predictors of VTE(31). In our cohort the length of VA ECMO cannulation was not associated with the rate of VTE. Our rate of DVT was significantly lower however, our cohort did not include V-V ECMO, and we only performed ultrasounds when clinically indicated. Despite the underreported rate of thrombosis eCPR patients are anticoagulated aggressively with more than half of them receiving antiplatelet therapy in addition to heparin given the burden of acute coronary syndrome as the cause of VT/VF arrest. Given the high rate of bleeding and low thrombotic risk in our eCPR population, further research should evaluate anticoagulation strategies to find the optimal approach that minimizes risk of bleeding without increasing thrombotic sequelae (26).

Given the retrospective nature of the study, some limitations are inevitable. While this represents one of the largest single center cohorts published, its small size may limit discrimination of small differences between groups. Despite performing adjusted analyses, we cannot account for potential confounding from variables that were not included in the model. The study was observational and change in anticoagulation management in response to bleeding, thrombotic events, or lab values was not protocolized or mandated and was not included in the analysis. Finally, despite using survival analysis and competing risk analysis, survival bias is unavoidable. Patients who die are no longer able to experience a bleeding or thrombotic event therefore our results should be interpreted with caution.

## Conclusions

In this large study of eCPR patients, 67.5 % of patients experienced major bleeding as defined by ELSO, while 23% of our cohort experienced thrombosis. Lower fibrinogen was associated with increased bleeding risk while the use of DAPT and lower antithrombin III levels on admission was associated with decreased thrombotic risk. Major bleeding was not associated with in-hospital mortality, but both thrombosis and major bleeding were associated with in-hospital morbidity with higher transfusion needs and longer mechanical ventilation days for those who bled. There is a need to assess optimal anticoagulation and antiplatelet regimens that minimize bleeding risk without increasing thrombotic consequences.

## Disclosure statement

The authors report no financial relationships or conflicts of interest regarding the content herein as well as no other relevant relationships to industry.

## Abbreviations

CPR: cardiopulmonary resuscitation
DAPT: Dual antiplatelet therapy
eCPR: Extracorporeal cardiopulmonary resuscitation
PCI: percutaneous coronary intervention
OHCA: Out-of-hospital cardiac arrest
VA-ECMO: Veno arterial extracorporeal membrane oxygenation
VT/VF: Ventricular tachycardia/Ventricular fibrillation

## Data Availability

Data is available upon request

## Notes

### Competing Interest Statement

The authors have declared no competing interest.

### Funding Statement

No funding

### Author Declarations

The study was reviewed and approved by the Institutional Review Board (IRB) at the UMN (IRB #1703M11301).

